# High Variability in Nicotine Analog Contents, Misleading Labeling, and Artificial Sweetener in New E-Cigarette Products Marketed as “FDA-Exempt”

**DOI:** 10.1101/2024.04.19.24306019

**Authors:** Hanno C. Erythropel, Sairam V. Jabba, Peter Silinski, Paul T. Anastas, Suchitra Krishnan-Sarin, Julie B. Zimmerman, Sven E. Jordt

**Affiliations:** Department of Chemical and Environmental Engineering, Yale University, New Haven, CT; Department of Chemistry, Duke University School of Medicine, Durham, NC; Department of Anesthesiology, Duke University School of Medicine, Durham, NC; Department of Psychiatry, Yale University School of Medicine, New Haven, CT

**Author notes:** Corresponding author: Sven E. Jordt, Ph.D., Associate Professor, Department of Anesthesiology, Duke University School of Medicine, 3 Genome Ct., Durham, NC 27710, USA. These authors contributed equally (Hanno C. Erythropel, Sairam V. Jabba).

## Abstract

The recent introduction of electronic cigarette products containing a synthetic nicotine analog, 6-methyl nicotine (6MN), challenges FDA’s tobacco regulatory authority. A similar strategy is pursued by vendors of recently introduced e-cigarette liquids containing nicotinamide (NA), marketed as ‘Nixotine’ or ‘Nixamide’. Compared to nicotine, 6MN is pharmacologically more potent at nicotinic receptors, and more toxic, raising concerns about increased addictiveness and adverse effects.

Here, combinations of gas chromatography, high performance liquid chromatography and mass spectrometry were used to determine nicotine analogs, flavor and sweetener contents of e-cigarette liquids of the brands “SpreeBar” and ECBlend “Nixotine” products. All SpreeBar products, labelled as containing 5% 6-methyl nicotine, contained only 0.61-0.64% 6-methylnicotine, while “Nixotine” samples contained 7-46% less of the declared nicotinamide contents. Although “Nixotine” product labels did not list 6MN as an ingredient, small amounts of 6-methyl nicotine were detected. All ‘SpreeBar’ samples contained the artificial sweetener neotame (0.20-0.86μg/mg).

Results identified significant discrepancies between declared and measured constituents of e-cigarette products containing nicotine alternatives. The discrepancy is misleading for consumers and raises concerns about production errors. ‘SpreeBar’ products also contained neotame, a high-intensity sweetener with high heat stability, likely increasing appeal to young and first-time users. Novel e-cigarette products with misleading labels containing nicotine analogs instead of nicotine on the US market is concerning and should be urgently addressed by lawmakers and regulators.

## INTRODUCTION

In March 2022 the Food and Drug Administration (FDA) received authorization to regulate tobacco products containing synthetic nicotine. The recent introduction of ‘SpreeBar’, an electronic cigarette (e-cigarette) containing a synthetic nicotine analog, 6-methyl nicotine (6MN) marketed as ‘Metatine’, challenges this authority (Figure). The vendor claims that FDA lacks authority over 6MN, since it differs chemically from nicotine, enabling marketing with youth-appealing flavors and exempt from tobacco product taxes.^1^ A similar strategy is pursued by vendors of recently introduced e-cigarette liquids containing nicotinamide (NA), marketed as ‘Nixotin’/’Nixodine’/’Nixamide’/’Nic-Safe’ claimed to be “carefully designed to target the same nicotinic acetylcholine receptors that traditional nicotine stimulates.” (Figure)^2^ ‘SpreeBar’ products are labeled as containing 5% 6MN, similar to nicotine content in Juul e-cigarettes (Figure). Compared to nicotine, 6MN is pharmacologically more potent at nicotinic receptors, and more toxic with a significantly lower LD_50_.^1^ This raises concerns about increased addictiveness and adverse effects of such nicotine replacement products. This study aimed to characterize and quantify constituents of ‘SpreeBar’ and ‘Nixotine’ products to assess product consistency and risk.

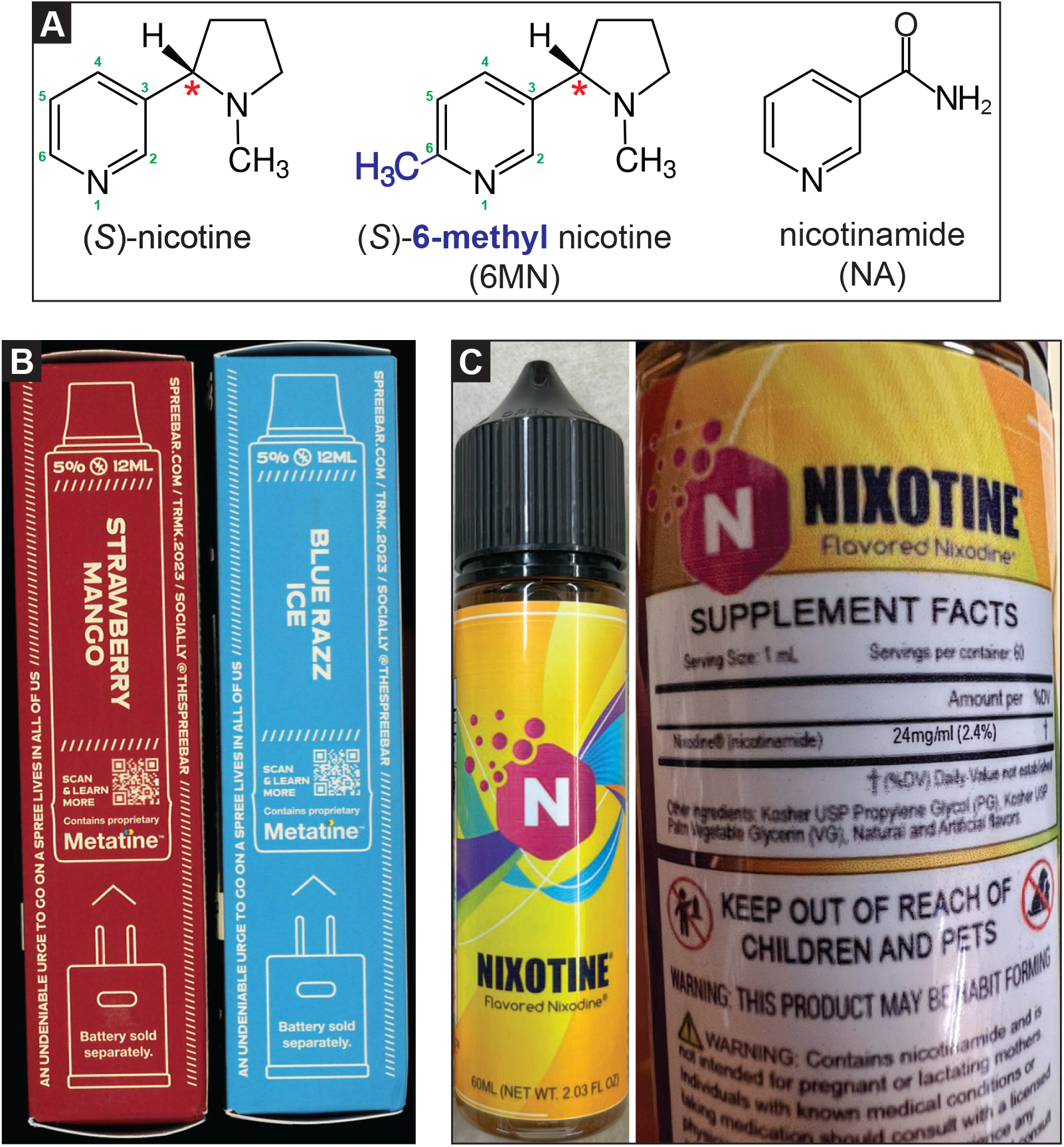
(**A**) Chemical structure of (S)-nicotine and nicotine analogues (S)-6-methyl nicotine and nicotinamide; the red star indicates the chiral center (**B**) Photo of side of cardboard packaging of 2 different ‘SpreeBar’ flavor pods, labelled “5%”, stating “Contains proprietary Metatine”, which is the trademark name for 6-methyl nicotine (bottom right); (**C**) Front and back of ‘Nixotine’ refill bottle, indicating only the presence of nicotinamide but not 6-methyl nicotine; indication of other ingredients (“Kosher USP Propylene Glycol (PG), Kosher USP Palm Vegetable Glycerin (VG), Natural and Artificial flavors”); and warning only about nicotinamide.

## METHODS

All available ‘SpreeBar’ flavors and 2 flavors of ‘Nixotine’ e-cigarette liquids at varying concentrations (Table) were purchased between 11/2023-2/2024 from online retailers (VaporFi, WestsideVapor; ECBlend, respectively). E-liquids were characterized via untargeted GC/MS analysis and all detected compounds were identified using the NIST database. Nicotine, nicotine analogs, synthetic cooling agents, artificial sweeteners, and other flavorants/additives were validated using commercial standards and quantified by GC/FID. Chiral HPLC/MS/MS was carried out to determine 6MN racemic purity, and non-chiral HPLC/MS/MS to quantify sweeteners using established methods^3^ (see Supplemental Materials).

**Table:** Labeled and quantified nicotine analog contents in ‘SpreeBar’ and ‘Nixotine’ products, calculated discrepancy, and measured solvent ratio (propylene glycol to glycerol), artificial sweetener neotame content, synthetic coolant WS-23 content, and whether other flavor compounds could be detected (see eAppendix for details). 6MN – 6-methyl nicotine; NA – nicotinamide; n.d. – not detected; PG – propylene glycol; GL – glycerol. Values shown as Mean (SE), all N ≥ 3.

| Product | Label nicotine alternative content | Measured nicotine alternative content [µg/mg] |  | Discrepancy in label and content (6MN for ‘Spree Bar’, NA for ‘Nixotine’) | Measured PG/VG ratio | Measured Neotame content in [µg/mg] | Measured WS-23 content in [µg/mg] | Other flavor compounds detected |
| --- | --- | --- | --- | --- | --- | --- | --- | --- |
|  |  | 6MN | NA |  |  |  |  |  |
| ‘SpreeBar’ products (labeled to contain 6MN) |  |  |  |  |  |  |  |  |
| Blood Orange Peach | 5%<br>= 50 µg/mg | 6.42 (0.07) | n.d. | -87% | 60/40 | 0.56 (0.03) | 4.44 (0.04) | Yes |
| Blue Razz Ice |  | 6.12 (0.05) | n.d. | -88% | 60/40 | 0.86 (0.12) | 16.39 (0.11) |  |
| Creamy Melon |  | 6.07 (0.17) | n.d. | -88% | 60/40 | 0.21 (0.01) | 5.74 (0.15) |  |
| Pineapple Coconut |  | 6.10 (0.04) | n.d. | -88% | 60/40 | 0.60 (0.03) | 4.47 (0.03) |  |
| Rainbowfruit |  | 6.23 (0.03) | n.d. | -88% | 60/40 | 0.65 (0.02) | 4.34 (0.07) |  |
| Strawberry Apple Melon |  | 6.41 (0.08) | n.d. | -87% | 60/40 | 0.20 (0.02) | 14.01 (0.18) |  |
| Strawberry Mango |  | 6.16 (0.11) | n.d. | -88% | 60/40 | 0.25 (0.01) | 9.81 (0.07) |  |
| Sweet Spearmint |  | 6.16 (0.07) | n.d. | -88% | 55/45 | 0.56 (0.01) | 16.18 (0.17) |  |
| Watermelon Grape |  | 6.28 (0.06) | n.d. | -87% | 60/40 | 0.51 (0.11) | 4.81 (0.05) |  |
| ‘Nixotine’ products (labeled to contain NA) |  |  |  |  |  |  |  |  |
| Unflavored | 0 mg/mL | n.d. | n.d. |  | 40/60 | n.d. | n.d. | No |
| Unflavored | 12 mg/mL | 0.09 (0.01) | 9.74 (0.05) | -7%<sup>a</sup> | 45/55 | n.d. | n.d. |  |
| Unflavored | 24 mg/mL | 0.13 (0.01) | 14.44 (0.00) | -31%<sup>a</sup> | 45/55 | n.d. | n.d. |  |
| Unflavored | 36 mg/mL | 0.15 (0.01) | 16.63 (0.06) | -46%<sup>a</sup> | 40/60 | n.d. | n.d. |  |
| Cherry | 0 mg/mL | n.d. | n.d. |  | 25/75 | n.d. | n.d. | Yes |
| Cherry | 12 mg/mL | 0.06 (0.01) | 8.32 (0.07) | -18%<sup>a</sup> | 30/70 | n.d. | n.d. |  |
| Cherry | 24 mg/mL | 0.10 (0.01) | 12.95 (0.04) | -35%<sup>a</sup> | 25/75 | n.d. | n.d. |  |
| Cherry | 36 mg/mL | 0.11 (0.02) | 16.47 (0.08) | -46%<sup>a</sup> | 30/70 | n.d. | n.d. |  |

<sup>a</sup> For ‘Nixotine’ products, µg/mg was converted to µg/mL using an estimated density of 1.15-1.2 g/mL based on measured PG/VG values (see eAppendix).

## RESULTS

No product contained nicotine. 6MN was detected in ‘SpreeBar’, and both 6MN and NA in ‘Nixotine’ products. However, ‘SpreeBar’ samples, labelled to contain 5% 6MN, had measured concentrations of only 0.61-0.64% 6MN (−87-88% less), while ‘Nixotine’ samples contained 7-46% less of the declared NA contents with discrepancies largest in the products with the highest labelled concentrations. Although ‘Nixotine’ product labels did not list 6MN as an ingredient, small amounts of 6MN were detected (Table). Chiral chromatography revealed that ‘SpreeBar’ contained only the (S)-6MN enantiomer. In addition to flavor-specific flavorants (Supplementary Table S1), all ‘SpreeBar’ samples contained the artificial sweetener neotame (0.20-0.86μg/mg) and the synthetic cooling agent WS-23 (4.34-16.39μg/mg; Table). ‘Nixotine’ products did neither contain sweeteners nor cooling agents.

## DISCUSSION

Results identified significant discrepancies between declared and measured constituents of e-cigarette products marketed as “PMTA-exempt”, claimed to be outside of FDA’s tobacco regulatory authority. While the significantly lower contents of 6MN (>87% lower) in ‘SpreeBar’ products may alleviate toxicological concerns, the discrepancy is misleading for consumers and raises concerns about production errors. ‘Nixotine’ products contained both NA and 6MN, with only the latter known to be a nicotinic receptor agonist.^1,4^ While the mixture, sold as ‘Nixodine’, may result in stimulant activity, the presence of 6MN is undeclared on the product label, misleading users and regulators.

Along with several youth-appealing flavorants, ‘SpreeBar’ products also contained neotame, a high-intensity sweetener, and WS-23, an odorless synthetic coolant. Neotame is 7,000-13,000x sweeter than sucrose and FDA-approved for use in food,^5^ has high heat stability, but has not been identified in US-marketed e-cigarettes to date. WS-23 is ubiquitous in disposable e-cigarettes.^6^ The presence of both compound classes is of particular concern as they increase appeal especially to young and first-time users, yet their inhalational safety remains unknown.

Taken together, the appearance of novel e-cigarette products with misleading labels containing nicotine analogs instead of nicotine on the US market is concerning and should be urgently addressed by lawmakers and regulators. FDA should be empowered to regulate them as tobacco products or designate them as drugs.

## Data Availability

All data produced in the present study are available upon reasonable request to the authors

## Author contributions

Drs Erythropel and Jabba contributed equally. Drs Erythropel, Jabba and Jordt had full access to all of the data in the study and take responsibility for the integrity of the data and the accuracy of the data analysis.

## Concept and design

Erythropel, Jabba, Zimmerman, Jordt.

## Acquisition, analysis, or interpretation of data

Jabba, Erythropel, Silinski, Krishnan-Sarin, Jordt.

## Drafting of the manuscript

Erythropel, Jabba, Jordt.

Critical revision of the manuscript for important intellectual content: Anastas, Zimmerman, Krishnan-Sarin, Jordt.

## Statistical analysis

Jabba, Erythropel, Jordt.

## Supervision

Jordt, Zimmerman.

## Disclaimer

The content is solely the responsibility of the authors and does not necessarily represent the views of the NIH or the FDA.

