## Supplemental Materials for "High Variability in Nicotine Analog Contents, Misleading Labeling, and Artificial Sweetener in New E-Cigarette Products Marketed as “FDA-Exempt”"

**Table S1: Flavorant compounds detected in tested “SpreeBar” and “Nixotine” products.**
**NA: nicotinamide**

| | Flavorant compounds identified by GC/MS (NIST database) |
|---|---|
| ‘SpreeBar’ products | |
| Blood Orange Peach | γ-Decalactone, ethyl butyrate, ethyl hexanoate, 3-hexenol, 3-hexenyl butyrate, hexyl acetate, isoamyl acetate, linalool, methyl octanoate, nerol, rheosmin, α-terpineol, WS-23[a] |
| Blue Razz Ice | Methyl benzoate, 2-methylbutyl 2-methylbutanoate, raspberry ketone, rheosmin, WS-23 |
| Creamy Melon | Ethyl butyrate, ethyl maltol, ethyl vanillin, ethylvanillin propylene glycol acetal, melonal, 6-nonenol, vanillin, vanillin propylene glycol acetal, WS-23 |
| Pineapple Coconut | Allyl hexanoate, γ-decalactone, ethyl butyrate, ethyl heptanoate, ethyl hexanoate, 3-hexenol, 3-hexen-1-ol acetate, maltol, WS-23 |
| Rainbowfruit | Benzyl alcohol, α-cumyl alcohol, ethyl butyrate, methyl anthranilate, WS-23 |
| Strawberry Apple Melon | Benzaldehyde propylene glycol acetal, γ-decalactone, ethyl butyrate, ethyl hexanoate, 3-hexenol, hexyl acetate, isoamyl 2-methyl butyrate, linalool, methyl hexanoate, methyl cinnamate, WS-23 |
| Strawberry Mango | Benzyl alcohol, γ-decalactone, ethyl butyrate, 3-hexenol, WS-23 |
| Sweet Spearmint | Carvone, eucalyptol, limonene, menthol, menthone, vanillin, WS-23 |
| Watermelon Grape | Benzyl acetate, benzyl alcohol, γ-decalactone, ethyl butyrate, 3-hexenol, menthyl acetate, methyl cinnamate, WS-23 |
| ‘Nixotine’ products | |
| Unflavored 0 mg/mL NA | none |
| Unflavored 12 mg/mL NA | |
| Unflavored 24 mg/mL NA | |
| Unflavored 36 mg/mL NA | |
| Cherry 0 mg/mL NA | Benzaldehyde, benzaldehyde propylene glycol acetal, benzaldehyde glycerol acetal |
| Cherry 12 mg/mL NA | |
| Cherry 24 mg/mL NA | |
| Cherry 36 mg/mL NA | |

[a] IUPAC name of WS-23: 2-Isopropyl-N,2,3-trimethylbutanamide; CAS # 51115-67-4

**Supplemental Methods**

**Chemicals & analytical standards**: Methanol, Water + 0.1% formic acid (FA), ACN + 0.1% FA, ammonium formate, propylene glycol, and glycerol were purchased >99% purity from Fisher Scientific (Waltham, MA). Nicotinamide and neotame at purities >99% and ammonium hydroxide as a 25% solution were purchased from Sigma-Aldrich, St. Louis, MO. Racemic (+/-) 6-methyl nicotine was purchased from Santa Cruz Biotechnology, Dallas, TX.

**GC/MS**: Gas chromatography (GC) coupled with mass spectrometry (MS) was carried out on a Clarus 580 GC coupled with SQ8S MS (PerkinElmer, Waltham, MA), fitted with a Elite-5MS column (length 60 m, id 0.25 mm, 0.25 µm film; PerkinElmer). Injection volume: 1 µL; split ratio: 10; program: 40 °C for 7 min; ramp 10 °C/min to 50 °C then hold for 20 min; ramp 10 °C/min to 310 °C then hold for 8 min; ramp 10 °C/min to 325 °C then hold for 11.5 min. Injector temperature: 300 °C; MS settings: ionization mode: electron impact ionization (EI+); m/z 30-620.

**GC/FID**: GC coupled with flame ionization detection (FID) was carried out on a GC-2010 (Shimadzu, Columbia, MD) fitted with a J&W DB-5 column (length 60 m, id 0.25 mm, 0.25 µm film; Agilent, Santa Clara, CA). Injection volume: 1 µL; split ratio: 300; program: 30 °C for 7 min; ramp 10 °C/min to 50 °C then hold for 20 min; ramp 10 °C/min to 310 °C then hold for 12 min. Injector temperature: 250 °C; detector temperature: 325 °C. Calibration curves for 6-methyl nicotine (6MN) and nicotinamide (NA) with ≥ 4 points were constructed and used to quantify 6MN in "SpreeBar" products and NA in "Nixotine" products. 6MN: Limit of quantification: 30 ug/mL; limit of detection: 10 ug/mL. NA: Limit of quantification: 35 ug/mL, limit of detection: 10 ug/mL.

**Chiral LC/MS/MS for 6MN**: Liquid chromatography (LC) coupled with MS/MS was carried out on a Vanquish LC coupled to a Quantis MS (Thermo Fisher Scientific). Chiral separation was achieved using a ChiralPAK AGP column (150 mm x 4 mm id, 5 µm particle size; Daïcel Corporation, West Chester, PA) using an isocratic mobile phase program consisting of 90:10 (v/v) of 30 mM ammonium formate with 0.3% $NH_4OH$ and methanol at 0.4 mL/min flow with a total runtime of 25 min at 25°C and an injection volume of 10 µL. MS/MS was carried out in heated electrospray ionization (H-ESI) mode at spray voltage: 3500 V (positive), 2500 V (negative); ion transfer tube temperature 300 °C; vaporizer temperature 350 °C; using selected reaction monitoring (SRM) mode for the following compounds:

| **Compound** | **Polarity** | **Precursor (m/z)** | **Product 1 (m/z)** (Quantifying) | **Product 2 (m/z)** (Confirming) | **Product 3 (m/z)** (Confirming) |
|---|---|---|---|---|---|
| 6-Methyl nicotine | Positive | 177.1 | 146.0 | 144.0 | 130.9 |
| Nicotine | Positive | 163.1 | 117.0 | 130.0 | 132.0 |

Absent optically pure standards, (S)-6MN was tentatively identified based on the relative retention time of pure (S)-nicotine, which elutes after (R)-nicotine in the described conditions. For 'SpreeBar' samples, only the second of the two peaks detected in the racemic (+-) 6MN was detected for all samples:

| | |
|---|---|
| 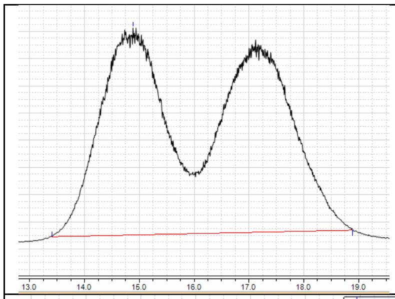 | SRM results for (+-) 6MN standard<br>(X-axis: time, Y-axis: counts) |
| 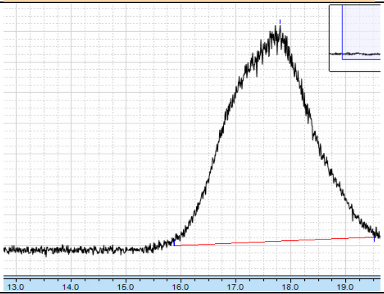 | Example SRM result, here for SpreeBar Watermelon Grape<br>(X-axis: time, Y-axis: counts) |

**LC/MS/MS for artificial sweeteners**: Liquid chromatography (LC) coupled with MS/MS was carried out on a Vanquish LC coupled to a Quantis MS (Thermo Fisher Scientific). Compound separation was achieved via an established procedure using an Accucore RP-MS column (50 mm x 2.1 mm id, 2.6 µm particle size, Thermo Fisher Scientific) using a binary mobile phase program of (A) water + 0.1% formic acid and (B) acetonitrile + 0.1% formic acid at 0.4 mL/min flow with a total runtime of 8 min at 40 °C and an injection volume of 10 µL. The gradient was as follows:

| Time (min) | A (%) | B (%) |
|---|---|---|
| 0 | 95 | 5 |
| 0.5 | 95 | 5 |
| 2.5 | 5 | 95 |
| 4.5 | 5 | 95 |
| 4.6 | 95 | 5 |

MS/MS was carried out in heated electrospray ionization (H-ESI) mode at spray voltage: 3500 V (positive), 2500 V (negative); ion transfer tube temperature 300 °C; vaporizer temperature 350 °C; using selected reaction monitoring (SRM) mode for the following compounds:

| Compound | Polarity | Precursor (m/z) | Product 1 (m/z) (Quantifying) | Product 2 (m/z) (Confirming) | Product 3 (m/z) (Confirming) |
|---|---|---|---|---|---|
| Acesulfame K | Negative | 161.9 | 81.9 | 77.9 | |
| Advantame | Negative | 443.1 | 425.0 | 243.9 | |
| Aspartame | Positive | 295.1 | 120.0 | 180.0 | 235.0 |
| Glucose | Negative | 76.9 | 60.1 | | |
| Glycyrrhizin | Negative | 821.4 | 351.0 | 113.0 | |
| Neotame | Positive | 380.2 | 172.0 | 320.0 | |
| Saccharin | Negative | 181.9 | 105.9 | 42.0 | |
| Sodium cyclamate | Negative | 177.9 | 79.9 | | |
| Sorbitol | Negative | 181 | 88.9 | 71.0 | 100.9 |
| Stevioside | Negative | 803.3 | 641.2 | 479.2 | |
| Sucralose | Negative | 397 | 361.0 | 359.0 | |
| Sucrose | Positive | 365 | 202.9 | 184.9 | |

Only neotame was detected in ‘SpreeBar’ samples and calibration curves for neotame with ≥ 4 points were constructed and used to quantify neotame in ‘SpreeBar’ products. Neotame: Limit of quantification: 5 ng/mL, limit of detection: 1 ng/mL.
